# Independent Genetic Effects of Glucagon-like Peptide-1 Receptor Locus on Body Mass Index and Type 2 Diabetes

**DOI:** 10.64898/2026.04.10.26350615

**Authors:** Chang Liu, Qin Hui, Gregorio V Linchangco, Amonae Dabbs-Brown, Jin J Zhou, Jacob Joseph, Peter D Reaven, Mary K Rhee, Luc Djousse, Kelly Cho, John Michael Gaziano, Peter WF Wilson, Lawrence S Phillips, VA Million Veteran Program, Yan V Sun

## Abstract

**Background:** The glucagon-like peptide-1 receptor (*GLP1R*) is a key regulator of glucose metabolism and appetite and a major therapeutic target for type 2 diabetes (T2D) and obesity. Genetic studies have implicated the *GLP1R* locus in both body mass index (BMI) and T2D, but it remains unclear whether their underlying genetic associations are the same.

**Methods:** We analyzed 431,107 participants of genetically inferred European ancestry from the Million Veteran Program. Within ± 500 kb of *GLP1R*, we performed locus-wide linear regression models for BMI and logistic regression models for T2D, adjusted for age, sex, and 10 principal components. We identified primary and secondary BMI sentinel variants using conditional analyses and evaluated their associations with T2D. Bayesian fine-mapping was used to construct credible sets of *GLP1R* locus for BMI and T2D.

**Results:** Conditioning on the primary sentinel variant rs12213929 (upstream of *GLP1R,* β = 0.11; 95% CI 0.09-0.14; p = 1.94×10^−17^), we identified a secondary variant (rs13216992, intron of *GLP1R*) independently associated with BMI (β = 0.10; 95% CI 0.07-0.13; p = 7.88×10^−14^). The two sentinel variants showed low linkage disequilibrium (r^2^ = 0.03). A two-variant allelic burden score (0-4; sum of the rs12213929 G-allele count and rs13216992 C-allele count) showed that participants with 4 risk alleles had 0.47 kg/m^2^ higher BMI than those with 0 risk alleles (95% CI 0.39-0.55; p < 2×10^−16^). Both variants were associated with higher T2D risk, but with distinct patterns after BMI adjustment: the rs12213929-T2D association persisted after adjustment for BMI (OR = 1.02; 95% CI 1.01-1.03; p = 0.0004), whereas the rs13216992-T2D association was fully attenuated (OR = 1.00; 95% CI 0.99-1.01; p = 0.68). Fine-mapping identified a compact 95% BMI credible set of 17 variants and a broader 95% T2D credible set of 42 variants, with all BMI credible variants contained within the T2D set.

**Conclusions:** The *GLP1R* locus harbors at least two independent BMI-associated variants that exhibit heterogeneous relationships with T2D: rs12213929 influences T2D risk partly through BMI-independent pathways, whereas rs13216992 appears to act predominantly via adiposity. These findings refine the genetic architecture at this key therapeutic target gene and provide a foundation for functional and pharmacogenomic studies to determine whether *GLP1R* variation can inform precision prevention and treatment of obesity and T2D.

## Introduction

The glucagon-like peptide-1 receptor gene (*GLP1R*) encodes a key receptor involved in glucose metabolism and appetite regulation ^1^. *GLP1R* is activated by the incretin hormone GLP-1 to stimulate insulin secretion and reduce food intake. GLP-1 receptor agonists (GLP1-RAs) such as semaglutide targets this pathway to treat for type 2 diabetes and obesity ^2–4^, and demonstrated protective effects of a broad range of cardiometabolic conditions ^5–10^. The introduction of an oral option expands beyond injections facilitates broader real-world uptake ^11–13^. In line with its physiological role, *GLP1R* has emerged as an important locus in human genetic studies of metabolic traits. Recent genome-wide association studies (GWAS) have identified *GLP1R* variants associated with body mass index (BMI) ^14^ and with risk of type 2 diabetes (T2D) ^15,16^, suggesting that genetic variation in this gene may influence both adiposity and glucose homeostasis, and contribute to obesity and diabetes risk.

Epidemiologic studies consistently show a strong association between higher BMI and increased T2D risk ^17^, while Mendelian randomization analyses support the causal effect of obesity on T2D ^18^. Genome-wide analyses further demonstrate a positive genetic correlation of 0.43 between BMI and T2D, consistent with substantial shared polygenic architecture between adiposity and diabetes risk ^19^. Together, these findings support the role of BMI as a key, partly genetically driven risk factor for T2D, while leaving room for BMI-independent genetic mechanisms that also influence diabetes susceptibility.

Despite *GLP1R’s* dual relevance to BMI and T2D, the independent genetic effects at this locus remain unclear. Because obesity is a major risk factor for T2D, it is challenging to determine whether *GLP1R* variants influence T2D directly or indirectly via BMI. To address this knowledge gap, we fine-mapped the *GLP1R* genomic region in a large, well-characterized cohort to identify and distinguish association signals for BMI and T2D. Our goal was to identify independent genetic associations with BMI and T2D in the region, and to evaluate whether distinct *GLP1R*-linked variants underlie these two traits. This fine-mapping approach allows us to elucidate the underlying genetic effects of the *GLP1R* locus and explore how allelic heterogeneity may differentially affect adiposity and diabetes risk.

## Methods

### Study population

We conducted all analyses in the Million Veteran Program (MVP), an ongoing mega-biobank within the U.S. Department of Veterans Affairs that links survey data, blood biospecimens, and genotypes to longitudinal electronic health records (EHRs) for more than one million U.S. veterans ^20^. Participants provided written informed consent for genetic research and EHR linkage. For the present analysis, we restricted to 431,107 MVP participants of genetically inferred European ancestry with available genotype data and BMI measurement and T2D phenotype information. participants assigned to the European ancestry group using the Harmonized Ancestry and Race/Ethnicity (HARE) algorithm, which is a machine learning based approach that integrates self-identified race/ethnicity with genetically inferred ancestry to generate harmonized ancestry groups in MVP ^21^.

### Genotyping, quality control, and imputation

Participants were genotyped using the custom MVP 1.0 Affymetrix Axiom array, which includes genome-wide tag SNPs and enriched content for cardiovascular and metabolic traits, as previously described ^22^. Standard sample- and variant-level quality control procedures were applied following the central MVP pipeline, including removal or quarantine of samples with low call rate, excess heterozygosity, relatedness, and sex misclassifications ^22^. Genotypes were imputed to the 1000 Genomes Project Phase 3 reference panel (GRCh37/hg19) using established pipelines ^22,23^. For downstream analyses, we additionally excluded variants with low minor allele frequency of <1% or showed extreme deviation from Hardy-Weinberg equilibrium with p < 1×10^−20^.

### Phenotype definitions

Body mass index was measured using height and weight measurements extracted from the VA EHR. For each participant, BMI was calculated as weight (kg) divided by height (m^2^). T2D status was defined using a validated EHR-based phenotyping algorithm that integrates diagnosis codes (ICD-9/ICD-10), antidiabetic medication prescriptions, and glycemic laboratory measures (fasting plasma glucose and HbA1c), consistent with prior MVP genetic analyses ^24^. All remaining participants without evidence of T2D were classified as non-T2D controls.

### Primary and secondary genetic associations with BMI in the *GLP1R* region

The primary sentinel SNP for BMI was identified among the 431,107 participants of European ancestry in the MVP. Within the *GLP1R* locus, which is ± 500 kb of the protein coding region, the strongest association with BMI was observed at the sentinel SNP rs12213929, located in the *DNAH8* gene upstream of the *GLP1R* coding region. This variant reached genome-wide significance in MVP in the linear regression model after adjustment for age, sex, and the top 10 principal components (PCs) of derived using the full genomics data.

We then performed conditional analysis to identify additional independent signals. Specifically, in the secondary BMI analysis, we repeated the locus-wide regression models adjusting for age, sex, PCs, and the primary sentinel SNP rs12213929 to search for residual association peaks. The secondary sentinel variant was defined as the SNP that remained significantly associated with BMI after conditioning on rs12213929 and showed a clear, distinct association peak, with the sentinel SNP of rs13216992.

In addition, we quantified genetic burden using the two independent BMI-associated variants. For each SNP, we coded genotype additively as the number of BMI-increasing alleles (allele G for rs12213929, and allele C for rs13216992). We then created a combined risk-allele count by summing the two allele counts, yielding a 5-level variable ranging from 0 to 4, with higher values indicating a greater number of risk alleles across the two loci. We evaluated the association of this combined score with continuous BMI using linear regression adjusted for age, sex, and the first 10 PCs.

### Association between BMI Sentinel SNPs and T2D

The associations between the primary and secondary BMI sentinel SNPs and T2D were assessed using logistic regression models. For the primary BMI sentinel SNP rs12213929, three sequential models were fitted: Model 1 adjusted for age, sex, and top 10 principal components; Model 2 further adjusted for BMI to evaluate whether the SNP-T2D association was independent of BMI; and Model 3 additionally included the secondary BMI sentinel SNP (rs13216992). For the secondary BMI sentinel SNP rs13216992, an analogous modeling strategy was applied, with Model 3 included the primary BMI sentinel SNP rs12213929 to assess whether its association with T2D was independent of the primary sentinel SNP.

To assess interaction between the two BMI sentinel SNPs and obesity status, we defined a binary indicator for obesity (BMI ≥30 kg/m^2^ vs <30 kg/m^2^). We then fit regression models for T2D status, adjusting for age, sex, 10 PCs. This analysis was conducted separately for rs12213929 and rs13216992.

### Regional fine-mapping and credible set construction

To characterize putative causal variants at *GLP1R* for BMI and T2D, we performed Bayesian fine-mapping and constructed credible sets separately for each trait using results of the locus-wide regression models adjusting for age, sex and top 10 PCs. We delineated credible causal sets by expanding the primary sentinel SNP for BMI to include proxy and putatively functional variants within a 500-kb window on either side using LDlinkR and the 1000 Genomes reference panel ^23,25^. Next, Bayesian fine-mapping with corrcoverage ^26–28^ was used to estimate variant-level posterior inclusion probability (PIP). Variants were then ranked by posterior probability, and 90% and 95% credible sets were defined as the smallest sets of variants whose cumulative PIPs summed to ≥0.90 and ≥0.95, respectively.

The locus-wide regression models were analyzed in PLINK ^29^. All other analyses were performed using R version 4.5.1 (https://www.R-project.org/).

## Results

Among 431,107 MVP participants of European ancestry, 105,609 (24.5%) had T2D (**Table 1**). The mean age of the overall cohort was 63.6 ± 13.7 years, and most participants were male (92.6%). Compared with participants without T2D, those with T2D were older (66.8 vs. 62.6 years) and had higher BMI (32.2 vs. 28.7 kg/m^2^). Genotype distributions at the two *GLP1R* BMI sentinel variants differed modestly but significantly between T2D cases and non-T2D controls (**Table 1**).

**Table 1.** Cohort Characteristics of the Million Veteran Program.

| Characteristics | Overall Cohort<br>N = 431107 | Type 2 Diabetes<br>N = 105609 | No Type 2<br>Diabetes<br>N = 325498 | P |
| --- | --- | --- | --- | --- |
| Age, years | 63.61 (13.72) | 66.82 (9.50) | 62.56 (14.68) | <0.001 |
| Male (%) | 399119 (92.6) | 101255 (95.9) | 297864 (91.5) | <0.001 |
| Body Mass Index, kg/m<sup>2</sup> | 29.58 (5.77) | 32.24 (6.27) | 28.72 (5.32) | <0.001 |
| BMI sentinel SNP1 rs12213929 Genotype (%) |  |  |  |  |
| GG | 176907 (41.0) | 43990 (41.7) | 132917 (40.8) | <0.001 |
| GA | 183036 (42.5) | 44501 (42.1) | 138535 (42.6) |  |
| AA | 45532 (10.6) | 10877 (10.3) | 34655 (10.6) |  |
| BMI sentinel SNP2 rs13216992 Genotype (%) |  |  |  |  |
| GG | 189875 (44.0) | 46261 (43.8) | 143614 (44.1) | 0.004 |
| CG | 189314 (43.9) | 46341 (43.9) | 142973 (43.9) |  |
| CC | 47800 (11.1) | 12021 (11.4) | 35779 (11.0) |  |

### Identification of independent BMI-associated variants at the *GLP1R* locus

Within the *GLP1R* locus, we identified two independent BMI-associated sentinel variants in MVP (**Table 2**, **Figures 1-2**). In the primary analysis adjusted for age, sex, and top 10 PCs, the strongest association with BMI was observed at rs12213929, located upstream of *GLP1R* in the DNAH8 region. Each copy of the G allele at rs12213929 was associated with a 0.11 kg/m² higher BMI (95% CI 0.09-0.14; p = 1.94 × 10^−17^).

**Figure 1.**
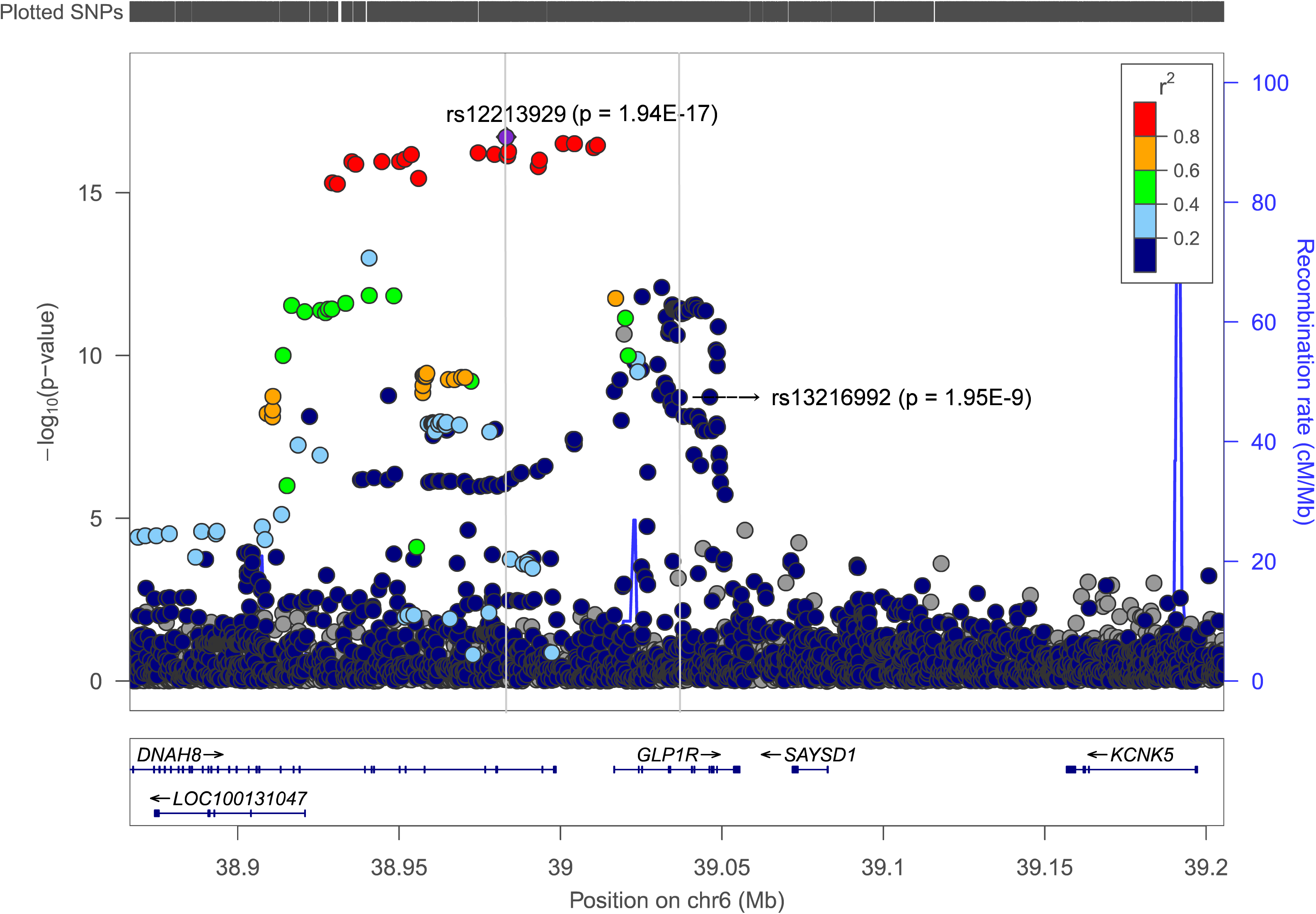
Regional plot of the association between the primary sentinel SNP rs12213929 and body mass index. Models adjusted for age, sex, the top 10 principal components. Two grey vertical lines indicate the locations of the primary sentinel SNP rs12213929 and the secondary sentinel SNP rs13216992.

**Figure 2.**
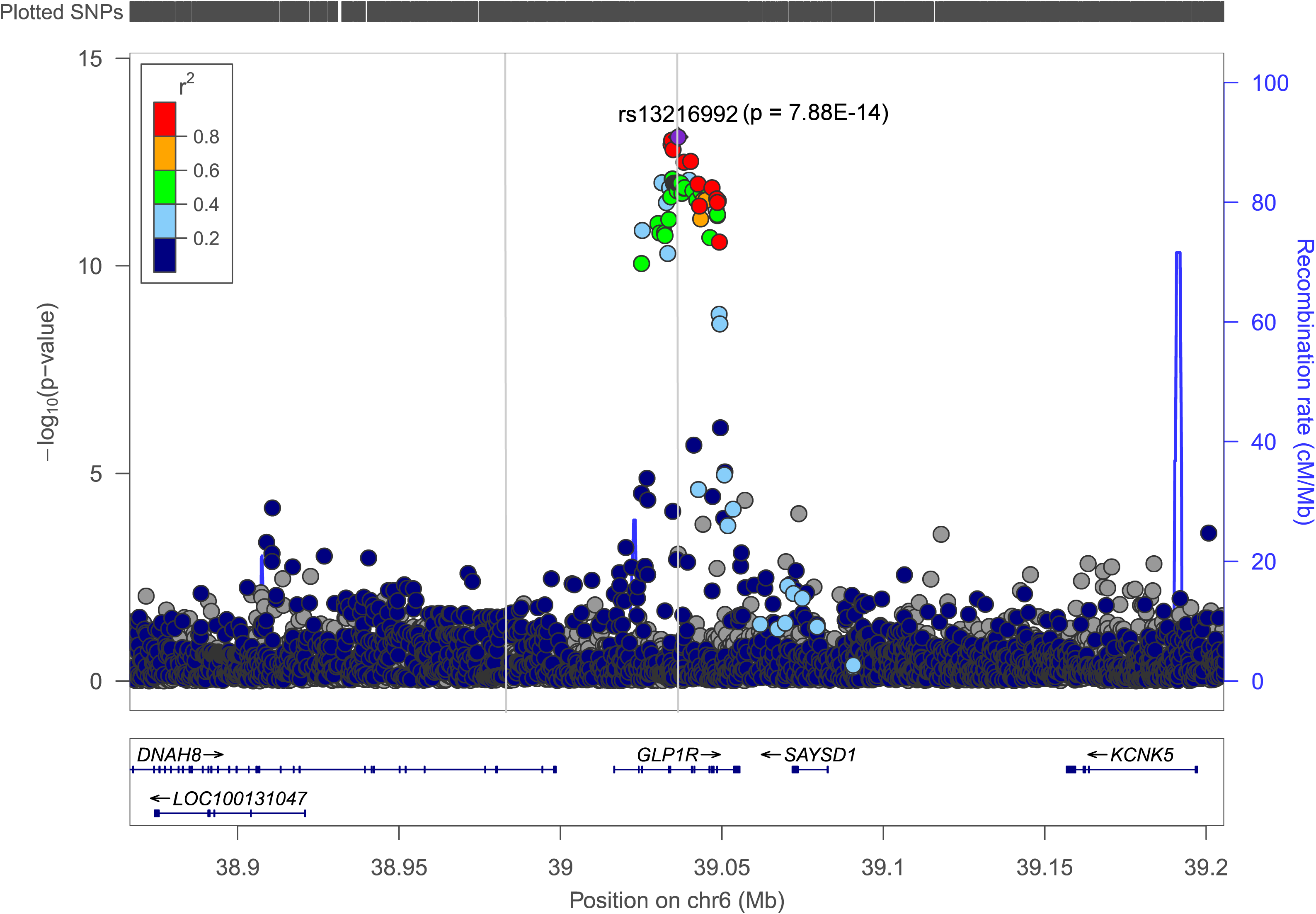
Regional plot of the association between the secondary sentinel SNP rs13216992 and body mass index. Models adjusted for age, sex, the top 10 principal components, and the primary SNP rs12213929. Two grey vertical lines indicate the locations of the primary sentinel SNP rs12213929 and the secondary sentinel SNP rs13216992.

**Table 2.** Association Between the *GLP1R* Variants and Body Mass Index in the Million Veteran Program.

| Variant | Model | Overall<br>N = 431107 |  |
| --- | --- | --- | --- |
|  |  | Beta<br>(95% CI) | P |
| **rs12213929**<br>(BMI sentinel SNP1,<br>effect allele G) | **Model 1: Age + Sex + PCs** | 0.11<br>(0.09, 0.14) | **1.94E-17** |
|  | **Model 2: Age + Sex + PCs + rs13216992** | 0.13<br>(0.10, 0.16) | **8.59E-22** |
| **rs13216992**<br>(BMI sentinel SNP2,<br>effect allele C) | **Model 1: Age + Sex + PCs** | 0.08<br>(0.05, 0.11) | **1.95E-9** |
|  | **Model 2: Age + Sex + PCs + rs12213929** | 0.10<br>(0.07, 0.13) | **7.88E-14** |

Conditional analysis adjusting for rs12213929 revealed a secondary independent signal at rs13216992. In models including age, sex, PCs, and rs12213929, the C allele at rs13216992 was significantly associated with higher BMI (β = 0.10; 95% CI 0.07-0.13; p = 7.88 × 10^−14^). Conversely, in models including rs13216992, the association of rs12213929 with BMI remained robust (β = 0.13; 95% CI 0.10-0.16; p = 8.59 × 10^−22^) (**Table 2**), supporting two independent BMI signals at the *GLP1R* locus. The two variants showed low linkage disequilibrium with r^2^ of 0.03.

When we further conditioned on both sentinel SNPs (rs12213929 and rs13216992) along with age, sex, and PCs, no additional variants in the *GLP1R* region reached genome-wide significance. The variant with the smallest residual p-value in this fully conditional model was rs147869249 (p = 2.17 × 10^−5^), which did not meet the genome-wide threshold, suggesting that rs12213929 and rs13216992 capture the major independent BMI associations at this locus.

In the linear regression of continuous BMI, the two-variant allelic burden score (0-4; sum of rs12213929 G-allele count and rs13216992 C-allele count) was positively associated with BMI. Each additional risk allele in the combined score was associated with 0.12 kg/m^2^ higher BMI (95% CI 0.10, 0.14; p < 2×10^−16^), after adjustment for age, sex, and 10 PCs, **Figure 3**.

**Figure 3.**
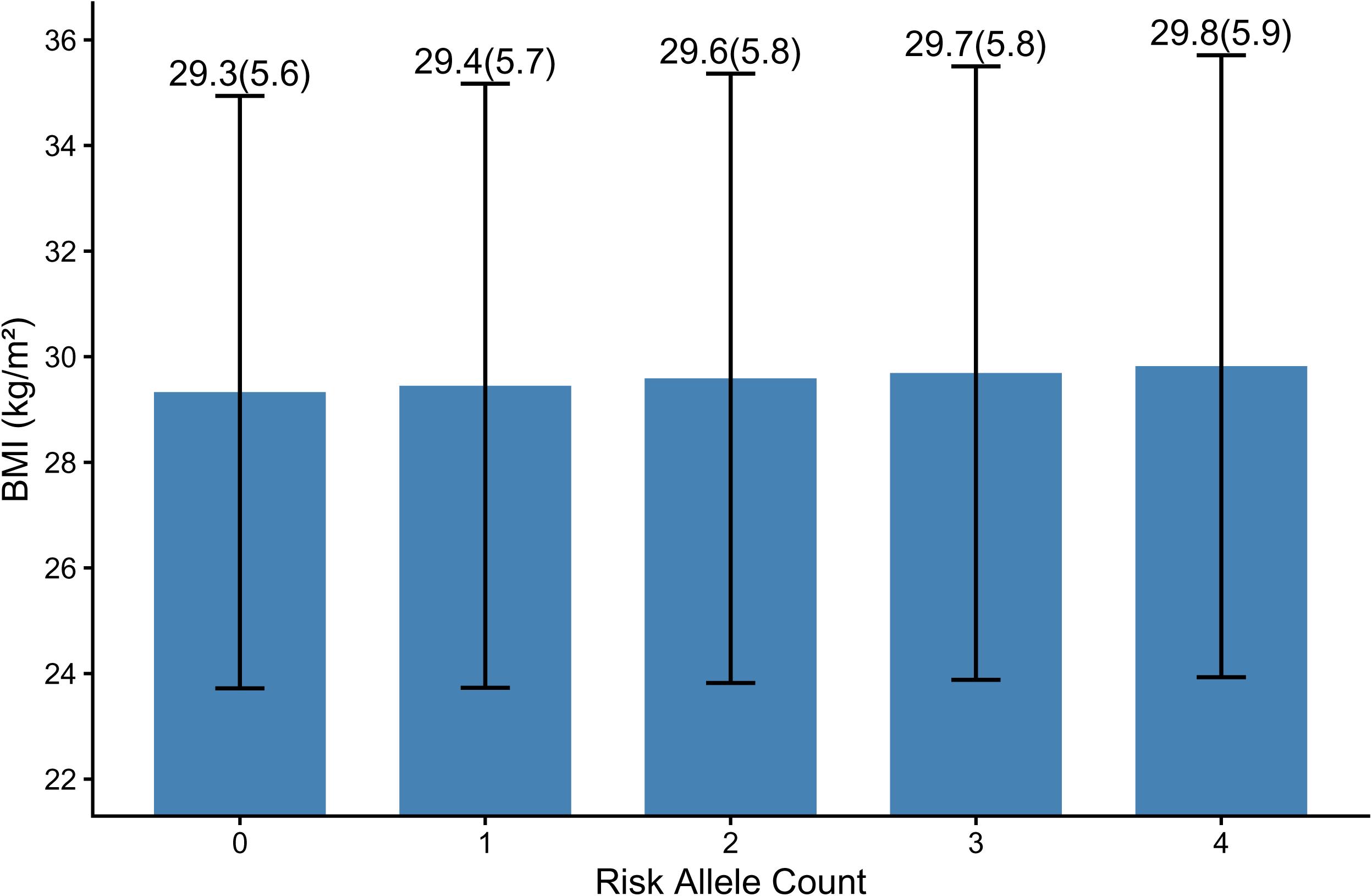
Mean body mass index by categories of the two-SNP allelic burden score, defined as the sum of the rs12213929 G allele count and the rs13216992 C allele count (range 0-4). Error bars indicate ± 1 standard deviation.

### Association of *GLP1R* BMI sentinel variants with type 2 diabetes

Both BMI sentinel variants were also associated with T2D risk in MVP, with distinct patterns regarding BMI adjustment (**Table 3**). For rs12213929, the G allele was associated with higher risk of T2D in models adjusted for age, sex, and PCs (OR = 1.03; 95% CI 1.02-1.04; p = 1.53 × 10^−9^). This association was only modestly attenuated after additional adjustment for BMI (OR = 1.02; 95% CI 1.01-1.03; p = 0.0004) and remained essentially unchanged when further adjusted for rs13216992 (OR = 1.02; 95% CI 1.01-1.03; p = 0.0003). These findings suggest that the rs12213929-T2D association is not fully mediated through BMI and is independent of the secondary BMI sentinel variant.

**Table 3.** Association Between the *GLP1R* Variants and Type 2 Diabetes in the Million Veteran Program.

| Cohort | Variant | Model | Estimates |  |
| --- | --- | --- | --- | --- |
|  |  |  | Odds Ratio<br>(95% CI) | P |
| Overall<br>N = 431107 | rs12213929<br>(BMI sentinel<br>SNP1,<br>effect allele G) | Model 1: Age + Sex + PCs | 1.03<br>(1.02, 1.04) | **1.53E-09** |
|  |  | Model 2: Age + Sex + PCs +BMI | 1.02<br>(1.01, 1.03) | **0.0004** |
|  |  | Model 3: Age + Sex + PCs +BMI +<br>rs13216992 | 1.02<br>(1.01, 1.03) | **0.0003** |
|  | rs13216992<br>(BMI sentinel<br>SNP2,<br>effect allele C) | Model 1: Age + Sex + PCs | 1.01<br>(1.00, 1.02) | **0.0278** |
|  |  | Model 2: Age + Sex + PCs +BMI | 1.00<br>(0.99, 1.01) | 0.6808 |
|  |  | Model 3: Age + Sex + PCs +BMI +<br>rs12213929 | 1.01<br>(0.99, 1.02) | 0.3175 |

BMI: Body Mass Index.

In contrast, the association of rs13216992 with T2D appeared to be largely mediated through BMI. In the model adjusting for age, sex and PCs, the C allele was associated with slightly higher T2D risk (OR = 1.01; 95% CI 1.00-1.02; p = 0.0278). However, this association was attenuated and became non-significant after adjustment for BMI (OR = 1.00; 95% CI 0.99-1.01; p = 0.6808). Additional adjustment for rs12213929 did not materially change the result (OR = 1.01; 95% CI 0.99-1.02; p = 0.3175), supporting BMI as the primary pathway linking rs13216992 to T2D.

In regression models of T2D status, there was no evidence of interaction between the two BMI sentinel SNPs and obesity (rs12213929: p = 0.3258; rs13216992: p = 0.0758).

### Fine-mapping and credible set analysis of the *GLP1R* locus

Fine-mapping of BMI associations at the *GLP1R* locus identified a relatively compact set of variants with high posterior probability of causality (**Table 4**). The 90% credible set consisted of 15 variants, with rs12213929 carrying the largest posterior probability (0.15). Extending to the 95% credible set added two additional variants, yielding a total of 17 variants that jointly accounted for ≥95% of the posterior probability for a causal BMI effect at the *GLP1R* locus.

**Table 4.** Credible-set Variants at the *GLP1R* Locus from Regional Association Analysis of BMI in the Million Veteran Program (adjusted for age, sex, and 10 principal components).

| rsID | Chr. | Position (hg19) | EA | NEA | EAF | Association with BMI |  | Credible Set Analysis |  |  |
| --- | --- | --- | --- | --- | --- | --- | --- | --- | --- | --- |
|  |  |  |  |  |  | Beta (95% CI) | P | Posterior Probability | In 90% CS | In 95% CS |
| rs12213929 | 6 | 38983183 | G | A | 0.66 | 0.1133 (0.0871, 0.1394) | 1.94E-17 | 0.15 | Yes | Yes |
| rs9380823 | 6 | 39000373 | G | T | 0.66 | 0.111 (0.0852, 0.1367) | 3.06E-17 | 0.10 | Yes | Yes |
| rs72857945 | 6 | 39003874 | A | G | 0.65 | 0.1096 (0.0841, 0.135) | 3.07E-17 | 0.10 | Yes | Yes |
| rs9380825 | 6 | 39010963 | G | A | 0.66 | 0.1114 (0.0855, 0.1373) | 3.43E-17 | 0.09 | Yes | Yes |
| rs9357297 | 6 | 39009846 | T | C | 0.66 | 0.1112 (0.0853, 0.1371) | 4.05E-17 | 0.07 | Yes | Yes |
| rs12214049 | 6 | 38983458 | G | C | 0.66 | 0.1106 (0.0848, 0.1365) | 5.34E-17 | 0.06 | Yes | Yes |
| rs1929902 | 6 | 38974055 | G | A | 0.66 | 0.111 (0.085, 0.137) | 5.87E-17 | 0.05 | Yes | Yes |
| rs1412265 | 6 | 38953335 | C | A | 0.67 | 0.1138 (0.0871, 0.1405) | 6.63E-17 | 0.05 | Yes | Yes |
| rs9357296 | 6 | 38979128 | A | G | 0.66 | 0.1108 (0.0848, 0.1368) | 6.53E-17 | 0.05 | Yes | Yes |
| rs12213825 | 6 | 38983182 | C | A | 0.66 | 0.1106 (0.0846, 0.1366) | 7.23E-17 | 0.04 | Yes | Yes |
| rs12213726 | 6 | 38982993 | C | T | 0.66 | 0.1106 (0.0846, 0.1366) | 7.25E-17 | 0.04 | Yes | Yes |
| rs9394561 | 6 | 38951317 | C | T | 0.66 | 0.1132 (0.0865, 0.1398) | 9.22E-17 | 0.03 | Yes | Yes |
| rs12202871 | 6 | 38993035 | T | C | 0.65 | 0.1091 (0.0834, 0.1349) | 9.85E-17 | 0.03 | Yes | Yes |
| rs1412263 | 6 | 38949692 | G | A | 0.67 | 0.1132 (0.0865, 0.14) | 1.08E-16 | 0.03 | Yes | Yes |
| rs1537230 | 6 | 38944196 | G | A | 0.67 | 0.1133 (0.0865, 0.1401) | 1.09E-16 | 0.03 | Yes | Yes |
| rs9380814 | 6 | 38934922 | G | A | 0.67 | 0.1134 (0.0866, 0.1402) | 1.10E-16 | 0.03 | No | Yes |
| rs9366987 | 6 | 38936096 | T | C | 0.67 | 0.1132 (0.0864, 0.14) | 1.31E-16 | 0.02 | No | Yes |

rsID, reference SNP identifier; Chr., chromosome; Position (hg19), base pair position based on human genome build 19; EA, effect allele; NEA, non-effect allele; EAF, effect allele frequency; CI, confidence interval; P, p-value; CS, credible set (90% CS, 90% credible set; 95% CS, 95% credible set).

Fine-mapping of T2D associations at *GLP1R* yielded a broader credible set (**Table 5**). The 90% credible set comprised 36 variants spanning the same regional block, and the 95% credible set included 42 variants.

**Table 5.** Credible-set Variants at the *GLP1R* Locus from Regional Association Analysis of Type 2 Diabetes in the Million Veteran Program (adjusted for age, sex, and 10 principal components).

| rsID | Chr. | Position (hg19) | EA | NEA | EAF | Association with Type 2 Diabetes |  | Credible Set Analysis |  |  |
| --- | --- | --- | --- | --- | --- | --- | --- | --- | --- | --- |
|  |  |  |  |  |  | OR (95% CI) | P | Posterior Probability | In 90% CS | In 95% CS |
| rs10305420 | 6 | 39016636 | C | T | 0.63 | 1.0339 (1.023, 1.0448) | 5.95E-10 | 0.08 | Yes | Yes |
| rs9357297 | 6 | 39009846 | T | C | 0.66 | 1.0341 (1.0231, 1.0452) | 7.53E-10 | 0.06 | Yes | Yes |
| rs9380825 | 6 | 39010963 | G | A | 0.66 | 1.034 (1.023, 1.0451) | 8.55E-10 | 0.05 | Yes | Yes |
| rs2268647 | 6 | 39043178 | T | C | 0.51 | 1.0323 (1.0219, 1.0429) | 9.41E-10 | 0.05 | Yes | Yes |
| rs72857945 | 6 | 39003874 | A | G | 0.65 | 1.0331 (1.0223, 1.044) | 1.13E-09 | 0.04 | Yes | Yes |
| rs4711569 | 6 | 38955590 | A | G | 0.66 | 1.0344 (1.0232, 1.0457) | 1.19E-09 | 0.04 | Yes | Yes |
| rs1929902 | 6 | 38974055 | G | A | 0.66 | 1.0338 (1.0228, 1.0449) | 1.23E-09 | 0.04 | Yes | Yes |
| rs9380823 | 6 | 39000373 | G | T | 0.66 | 1.0335 (1.0226, 1.0445) | 1.22E-09 | 0.04 | Yes | Yes |
| rs12202871 | 6 | 38993035 | T | C | 0.65 | 1.0334 (1.0225, 1.0445) | 1.26E-09 | 0.04 | Yes | Yes |
| rs9357296 | 6 | 38979128 | A | G | 0.66 | 1.0337 (1.0227, 1.0449) | 1.29E-09 | 0.04 | Yes | Yes |
| rs12214049 | 6 | 38983458 | G | C | 0.66 | 1.0336 (1.0226, 1.0446) | 1.35E-09 | 0.03 | Yes | Yes |
| rs12213726 | 6 | 38982993 | C | T | 0.66 | 1.0337 (1.0226, 1.0448) | 1.41E-09 | 0.03 | Yes | Yes |
| rs12213825 | 6 | 38983182 | C | A | 0.66 | 1.0337 (1.0226, 1.0448) | 1.41E-09 | 0.03 | Yes | Yes |
| rs12213929 | 6 | 38983183 | G | A | 0.66 | 1.0338 (1.0227, 1.045) | 1.53E-09 | 0.03 | Yes | Yes |
| rs1412265 | 6 | 38953335 | C | A | 0.67 | 1.0344 (1.023, 1.0458) | 1.83E-09 | 0.03 | Yes | Yes |
| rs1537230 | 6 | 38944196 | G | A | 0.67 | 1.0343 (1.0229, 1.0458) | 2.15E-09 | 0.02 | Yes | Yes |
| rs9394561 | 6 | 38951317 | C | T | 0.66 | 1.0341 (1.0228, 1.0456) | 2.18E-09 | 0.02 | Yes | Yes |
| rs1412263 | 6 | 38949692 | G | A | 0.67 | 1.0342 (1.0229, 1.0457) | 2.28E-09 | 0.02 | Yes | Yes |
| rs9380814 | 6 | 38934922 | G | A | 0.67 | 1.0343 (1.0229, 1.0458) | 2.30E-09 | 0.02 | Yes | Yes |
| rs9366987 | 6 | 38936096 | T | C | 0.67 | 1.0343 (1.0229, 1.0458) | 2.38E-09 | 0.02 | Yes | Yes |
| rs72853773 | 6 | 38928910 | A | G | 0.67 | 1.035 (1.0233, 1.0468) | 2.87E-09 | 0.02 | Yes | Yes |
| rs9380812 | 6 | 38930402 | C | T | 0.67 | 1.035 (1.0233, 1.0468) | 2.99E-09 | 0.02 | Yes | Yes |
| rs10305442 | 6 | 39024959 | G | A | 0.52 | 1.0322 (1.0215, 1.0431) | 2.92E-09 | 0.02 | Yes | Yes |
| rs2281342 | 6 | 38992668 | T | C | 0.66 | 1.0328 (1.0218, 1.0439) | 3.27E-09 | 0.01 | Yes | Yes |
| rs4711575 | 6 | 39038022 | A | G | 0.51 | 1.0307 (1.0204, 1.0412) | 4.16E-09 | 0.01 | Yes | Yes |
| rs4711573 | 6 | 39037877 | A | G | 0.51 | 1.0307 (1.0203, 1.0411) | 4.45E-09 | 0.01 | Yes | Yes |
| rs12214488 | 6 | 39036233 | G | A | 0.51 | 1.0303 (1.02, 1.0406) | 5.00E-09 | 0.01 | Yes | Yes |
| rs12201075 | 6 | 39036070 | A | G | 0.51 | 1.0302 (1.02, 1.0406) | 5.07E-09 | 0.01 | Yes | Yes |
| rs9296283 | 6 | 39034996 | A | G | 0.51 | 1.0301 (1.0199, 1.0404) | 5.12E-09 | 0.01 | Yes | Yes |
| rs7765879 | 6 | 39035521 | T | C | 0.51 | 1.0302 (1.02, 1.0406) | 5.23E-09 | 0.01 | Yes | Yes |
| rs7341358 | 6 | 39036470 | A | G | 0.51 | 1.0302 (1.02, 1.0406) | 5.23E-09 | 0.01 | Yes | Yes |
| rs9357298 | 6 | 39036834 | T | A | 0.51 | 1.0302 (1.02, 1.0406) | 5.23E-09 | 0.01 | Yes | Yes |
| rs7766068 | 6 | 39035647 | T | C | 0.51 | 1.0302 (1.02, 1.0406) | 5.23E-09 | 0.01 | Yes | Yes |
| rs7341356 | 6 | 39036345 | G | A | 0.51 | 1.0302 (1.02, 1.0406) | 5.26E-09 | 0.01 | Yes | Yes |
| rs7766275 | 6 | 39035836 | C | A | 0.51 | 1.0302 (1.02, 1.0405) | 5.32E-09 | 0.01 | Yes | Yes |
| rs7341360 | 6 | 39036579 | A | G | 0.51 | 1.0302 (1.02, 1.0405) | 5.29E-09 | 0.01 | Yes | Yes |
| rs7766228 | 6 | 39035713 | T | C | 0.51 | 1.0302 (1.02, 1.0405) | 5.32E-09 | 0.01 | No | Yes |
| rs9296282 | 6 | 39034734 | T | C | 0.51 | 1.0302 (1.0199, 1.0405) | 5.42E-09 | 0.01 | No | Yes |
| rs9394573 | 6 | 39037194 | A | G | 0.51 | 1.0302 (1.0199, 1.0405) | 5.63E-09 | 0.01 | No | Yes |
| rs9296281 | 6 | 39034636 | A | T | 0.51 | 1.0301 (1.0199, 1.0405) | 5.63E-09 | 0.01 | No | Yes |
| rs7765641 | 6 | 39035250 | A | G | 0.51 | 1.0301 (1.0199, 1.0405) | 5.65E-09 | 0.01 | No | Yes |
| rs9296284 | 6 | 39035132 | T | C | 0.51 | 1.0301 (1.0199, 1.0404) | 5.72E-09 | 0.01 | No | Yes |

rsID, reference SNP identifier; Chr., chromosome; Position (hg19), base pair position based on human genome build 19; EA, effect allele; NEA, non-effect allele; EAF, effect allele frequency; OR, odds ratio; CI, confidence interval; P, p-value; CS, credible set (90% CS, 90% credible set; 95% CS, 95% credible set).

Notably, all 17 variants in the 95% BMI credible set were also included in the 95% T2D credible set (**Tables 4-5**), indicating substantial overlap of putatively causal variants for BMI and T2D at the *GLP1R* locus.

## Discussion

In this genetic study of the *GLP1R* locus, we identified two independent genome-wide significant associations with BMI and demonstrated that they have distinct relationships with T2D. The primary upstream variant rs12213929 showed strong association with BMI and remained significantly associated with T2D after adjustment for BMI, whereas the secondary variant rs13216992 was associated with both higher BMI and higher T2D risk, but its T2D association was fully attenuated after BMI adjustment. Fine-mapping revealed that BMI and T2D share highly overlapping 95% credible sets at the *GLP1R* locus, suggesting largely shared causal variants for adiposity and diabetes within this region, while our conditional analyses indicate heterogeneity of individual variants act through BMI versus BMI-independent pathways.

These findings extend prior work implicating *GLP1R* in metabolic regulation and T2D susceptibility. GLP1-RAs are now widely used for T2D and obesity treatment ^3,30–32^. Genetic variation located in *GLP1R* region has been associated with glycemic traits and diabetes risk ^2,16,33–35^. Our results add to this literature by focusing on a regional fine-mapping of *GLP1R* and demonstrating that multiple independent variants in the locus contribute to BMI variation, while at least one of these variants (rs12213929) exerts a BMI-independent effect on T2D risk.

Our findings also align with accumulating evidence that *GLP1R* variation may influence response to GLP1-RA therapies. Pharmacogenomic studies have identified genetic variants, including in or near *GLP1R*, that modify glycemic response to GLP1-RAs ^36–38^. Our results suggest that allelic heterogeneity at the *GLP1R* locus may influence metabolic phenotypes through at least two partially distinct mechanisms: one that primarily acts through adiposity and another that exerts more direct effects on glucose homeostasis independent of BMI. These observations raise the hypothesis that individuals who carry *GLP1R* variants acting predominantly through BMI versus BMI-independent glycemic pathways may derive differential benefit from GLP-1 based therapies, with potentially distinct profiles of weight loss versus glucose lowering. If confirmed, such locus-specific heterogeneity could help identify subgroups of patients, with and without obesity, who are more likely to respond favorably to GLP1-RAs for prevention or treatment of T2D.

This study has several strengths. We leveraged individual-level data from a very large, well-characterized cohort of MVP, and we focused on a drug target gene with strong prior evidence for involvement in both BMI and T2D, allowing a detailed dissection of locus-specific architecture rather than a purely discovery-driven genome-wide screen. Several limitations should also be considered. Our analyses were restricted to participants of European ancestry, in which the genome-wide significant genetic association with BMI was originally reported ^34,39^. As a result, the identified sentinel variants and credible sets may not generalize fully to other ancestries, where differences in allele frequency and LD structure might refine or shift the set of putative causal variants. Future studies should directly evaluate *GLP1R* expression and downstream signaling in relevant tissues, and assess whether the sentinel variants we identified are associated with differential pharmacologic response to GLP-1 receptor agonists.

## Conclusion

In summary, we show that the *GLP1R* locus harbors at least two independent BMI-associated variants, and that these variants exhibit distinct patterns of association with T2D. The primary variant rs12213929 influences both BMI and T2D, with a residual BMI-independent effect on diabetes risk, whereas rs13216992 appears to act predominantly through BMI. These findings refine our understanding of the genetic architecture at this key therapeutic locus and provide a foundation for future functional and pharmacogenomic studies aimed at leveraging *GLP1R* variation for precision prevention and treatment of obesity and T2D.

## Acknowledgement

This research is based on data from the Million Veteran Program, Office of Research and Development, Veterans Health Administration, and was supported by MVP000 as well as award # MVP065 (Y.V.S.) as well as awards CX001737, BX005831, BX004821. This publication does not represent the views of the Department of Veteran Affairs or the United States Government. This research has also been supported in part by National Institutes of Health (NIH) grant P01 HL154996.

## Data Availability

Due to US Department of Veterans Affairs (VA) regulations and our ethics agreements, the analytic datasets used for this study are not permitted to leave the Million Veteran Program (MVP) research environment and VA firewall. This limitation is consistent with other MVP studies based on VA data. However, the MVP data are made available to researchers with an approved VA and MVP study protocol.

## Notes

### Competing Interest Statement

The authors have declared no competing interest.

### Author Declarations

The VA Central Institutional Review Board approved this study. All participants gave informed consent.

